# Long-read sequencing reveals hidden structural complexity within Alzheimer’s disease-associated GWAS loci

**DOI:** 10.64898/2026.03.16.26348487

**Authors:** Niccolò Tesi, Alex Salazar, Gerard Bouland, Daniel Alvarez, Yaran Zhang, Lydian Knoop, Natasja M. van Schoor, Martijn Huisman, Sanduni Wijesekera, Jana Krizova, Betty M. Tijms, Everard G.B. Vijverberg, Marc Hulsman, Sven van der Lee, Marcel Reinders, Henne Holstege

## Abstract

Genome-wide association studies (GWAS) typically represent Alzheimer’s disease (AD) risk loci by a single lead SNP, although the lead SNP may capture only part of the genetic variation at a locus. We asked what genetic architectures underlie AD GWAS associations and how much information is obscured when a locus is reduced to a single lead SNP.

We integrated AD GWAS summary statistics with long-read whole-genome sequencing of 493 individuals, phased DNA methylation, chromatin annotations, and structural variant (SV) imputation. Across 98 established AD loci, linkage disequilibrium (LD)-based clumping and conditional analysis identified 280 independent AD-haplotypes, of which 207 (74%) harbored linked structural variants, yielding 2,000 haplotype-linked SVs comprising intronic transposable elements and multiallelic tandem repeats within regulatory regions.

Integration of methylation, chromatin annotations, and genomic context prioritized 52 AD haplotypes with candidate regulatory SVs, revealing that lead SNPs often capture only part of the underlying structural and regulatory architecture. At the *SHARPIN/PLEC* locus, long-read fine-mapping resolved an independent regulatory *PLEC* haplotype carrying a CpG-rich tandem repeat associated with enhancer methylation and reduced microglial expression. Extending these findings to population scale, we imputed 83% of haplotype-linked SVs into SNP-genotyped cohorts (median imputation R^2^ = 0.76). In exploratory model comparisons, lead-SNP-only models were favored at just 10% of loci, whereas models incorporating multiple haplotypes, SVs, or both were favored at the remaining 90%.

Our findings show that AD GWAS loci frequently comprise structurally and regulatorily heterogeneous haplotypes that cannot be fully interpreted through the lead SNP alone. Rather than the lead SNP, the haplotype represents the fundamental unit for resolving the biological mechanisms driving AD genetic risk.

## Introduction

Alzheimer’s Disease (AD) is the most common form of dementia [1] and has a strong genetic component, with twin studies estimating its heritability at 60-80% [2]. Genome-wide association studies (GWAS) have identified 98 regions (loci) associated with a modest effect on AD risk [3, 4]. These associations are generally represented by the lead single-nucleotide polymorphism (SNP), although the lead SNP is not necessarily a functional variant. Instead, it may serve as a marker for a broader haplotype comprising multiple variants in linkage disequilibrium (LD) that collectively define the underlying genetic and regulatory architecture.

Here, we hypothesized that many AD GWAS signals are better understood as associations with haplotypes rather than with individual SNPs. A single GWAS locus can contain multiple conditionally independent association signals, each represented by a different lead SNP and embedded within a distinct haplotypic background. Furthermore, individuals carrying the same lead-SNP allele may still differ in the structural alleles and local haplotypic configurations present on that background. As a result, a single AD GWAS locus may harbor multiple disease-associated haplotypes, each defined by a unique combination of regulatory variants and structural variation. These distinct haplotypes may influence disease through different molecular mechanisms that cannot be captured by the lead SNP alone.

Structural variants (SVs), including transposable-element insertions (TEs) and tandem repeats (TRs), may be important components of these haplotypes. SVs represent a major source of genetic variation in the human genome [5] and are often associated with distinct local epigenetic and regulatory configurations, including DNA-methylation patterns, chromatin states and gene expression profiles [6–11]. Because many SVs are repetitive, multiallelic or located within complex genomic regions, they are poorly captured by SNP arrays and remain difficult to resolve using short-read sequencing. Consequently, a biallelic lead SNP may collapse multiple structurally and functionally distinct haplotypes into the same SNP-defined genotype class.

The importance of hidden structural variation is well established in neurological disease. Repeat expansions and other SVs have been implicated in various neurological diseases including Huntington’s disease, fragile X syndrome, and several forms of ataxia [12–14]. In AD, several observations suggest that structural variation contributes to the disease associations of risk loci [15–17]. A notable example is the *TMEM106B* locus, where we and others recently identified an AluYb8 retrotransposon embedded within an AD-associated haplotype [3, 18]. This transposable element co-segregates with the GWAS risk haplotype and is associated with local methylation differences, supporting a model in which the GWAS SNP acts primarily as a proxy for a potentially regulatory SV [18]. Whether this represents an isolated locus-specific finding or a broader feature of AD genetic architecture has not been systematically investigated.

Long-read sequencing now makes it possible to address this question. It enables the resolution of repetitive and multiallelic SVs while simultaneously providing phased genetic and DNA-methylation information [19, 20]. This offers an opportunity to move beyond lead-SNP annotation and directly characterize the structural and epigenetic composition of association-bearing haplotypes.

We therefore asked what genetic architectures underlie AD GWAS associations and how much information is obscured when a locus is reduced to a single lead SNP. We integrated AD GWAS summary statistics with long-read whole-genome sequencing, phased methylation profiles, chromatin annotations and SV imputation. We first decomposed established AD loci into conditionally independent association signals and characterized the haplotypic backgrounds carrying these signals. We then identified SVs linked to these haplotypes in 493 long-read sequenced genomes, and prioritized candidate regulatory SVs by integrating linkage, allele-specific methylation, genomic localization, and chromatin context. Finally, we evaluated whether these structurally resolved haplotypes could be imputed into larger SNP-genotyped cohorts and whether SV-aware models provided a more complete representation of AD association signals at population scale.

## Results

### AD GWAS loci frequently harbor multiple independent disease-associated haplotypes

We extracted summary statistics for 118 variants across 98 previously reported AD-associated loci, together with all SNPs located within 250 kilobases (kbp) of each locus (*Figure 1A-B, Table S1*) [4]. LD-based clumping identified 532 candidate haplotypes across all 98 loci showing suggestive association with AD (p<5×10^-5^, *Figure 1C, Table S2*, and Supplementary results). For each signal, the single most significantly associated SNP in the GWAS was retained as its representative marker.

**Figure 1:**
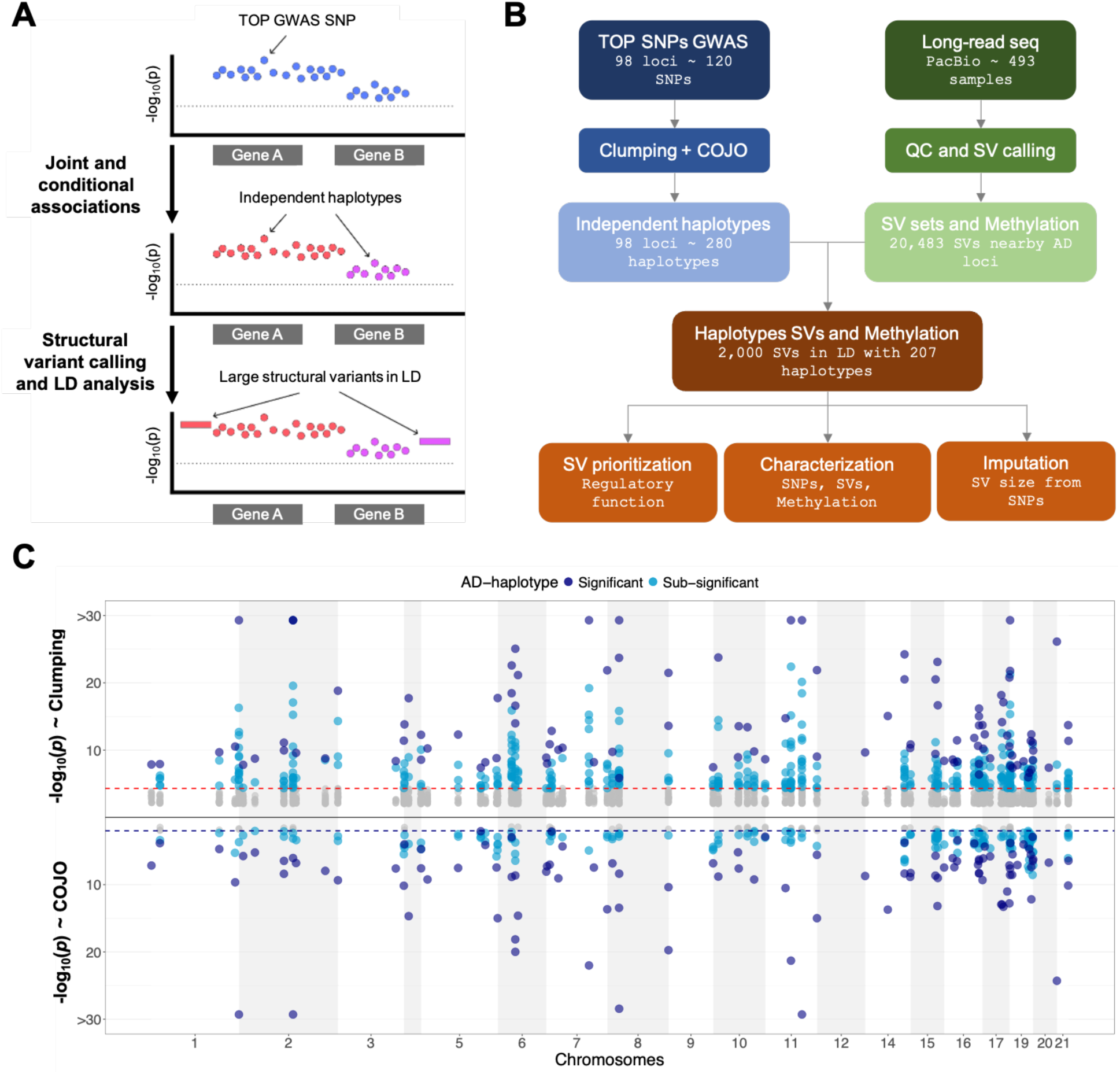
Analysis workflow and identification of AD-haplotypes. **A.** Schematic of the lead-SNP versus haplotype interpretation of a GWAS locus, illustrating that linked SNPs and structural variants on the same haplotype may underlie the association, and that multiple independent haplotypes can co-exist within a single locus. **B.** Workflow overview. AD-associated haplotypes were identified by clumping and conditional and joint association analysis of GWAS summary statistics. SVs and methylation signatures were genotyped from long-read sequencing of 493 Dutch individuals, and SVs were linked to haplotypes for prioritization, characterization, and imputation. **C.** Miami plot showing haplotypes after clumping and conditional analysis. Navy dots indicate known haplotypes associated with AD at genome-wide significance. Light blue dots indicate the sub-significant clumped and conditionally independent haplotypes. The red dashed line indicates the clumping threshold (p<5e-5), and the blue dashed line indicates the conditional analysis threshold used after multiple testing correction (False Discovery Rate p-value<1%).

We then used conditional and joint association (COJO) analysis to distinguish signals that remained associated with AD after accounting for local LD structure. In total, 280 conditionally independent signals remained significantly associated with AD after multiple testing correction (FDR<1%, *Figure 1C, Table S3*, and Supplementary results). For brevity, we refer to these conditionally independent, SNP-tagged signals as *AD-haplotypes*. Most loci harbored multiple independently associated AD-haplotypes, with *KCNN4, PLCG2, ABCA7*, and *ADAM10* containing at least 10, 10, 9, and 8 signals, respectively. These findings show that an AD GWAS locus is rarely represented adequately by a single lead SNP or a single associated genetic background. Instead, many loci comprise several independently associated haplotypes that may carry distinct variants and act through different molecular mechanisms. We therefore used long-read sequencing to resolve structural variation embedded within these AD-haplotypes.

### Long-read sequencing reveals extensive structural heterogeneity within AD-associated regions

PacBio Sequel II whole-genome sequencing of peripheral blood was generated for 245 AD patients from the Amsterdam Dementia Cohort (mean age 67.11, IQR=12, 70% females) [21] and 248 cognitively healthy centenarians from the 100-plus Study cohort (mean age 101.2, IQR=2, 70% females, *Figure 2A*) [22]. Across all individuals, median sequencing coverage including both HiFi and non-HiFi (read quality >0.85) was 25.7x and median read length was 14,616 bp (*Figure 2B*).

**Figure 2:**
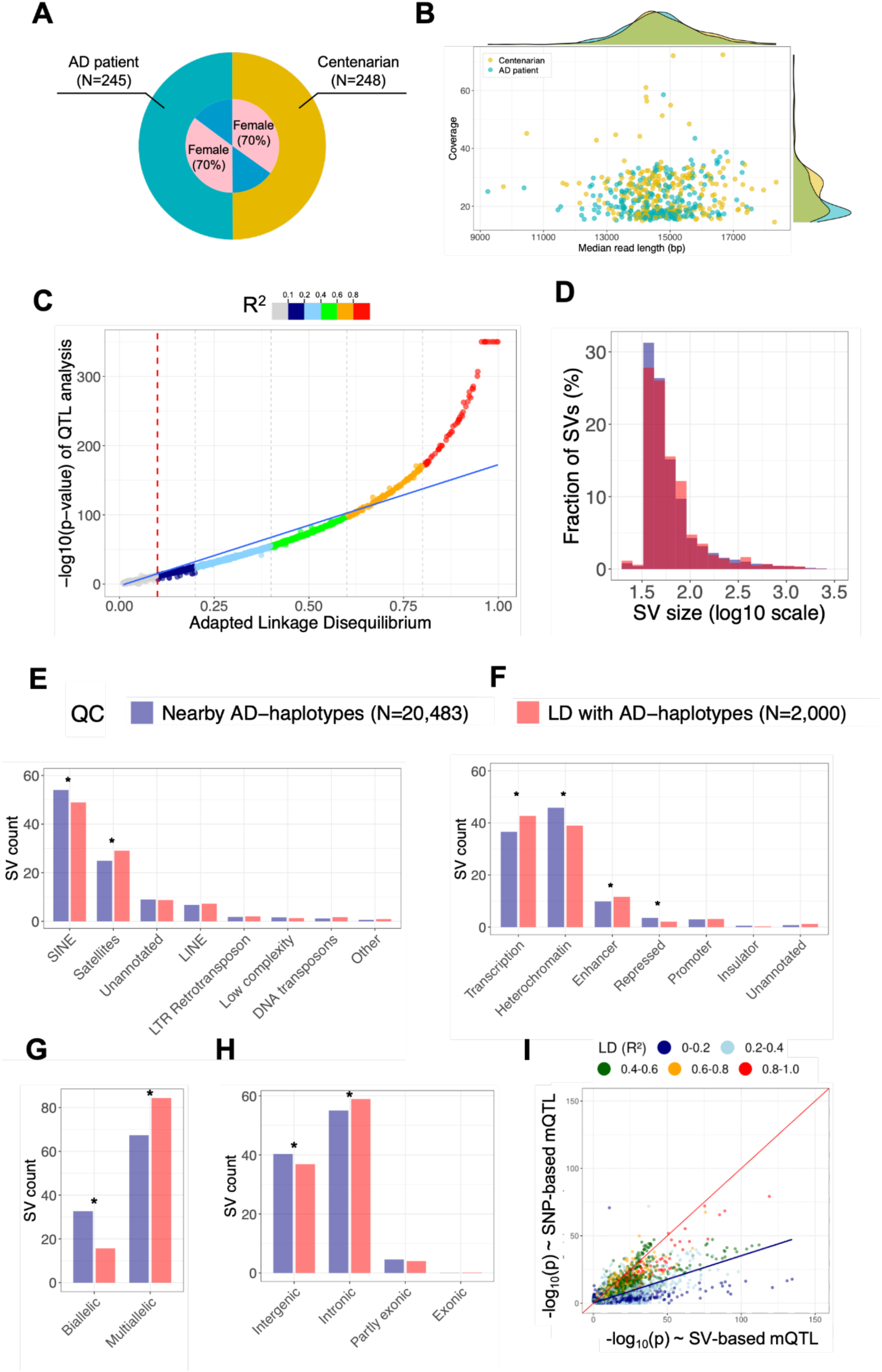
Long-read sequencing and characterization of SVs. Throughout panels D–H, blue indicates the full set of 20,483 variable SVs nearby AD-haplotypes, and red the subset of 2,000 SVs linked to AD haplotypes. **A.** Cohort composition: cognitively healthy centenarians (N=248) and AD patients (N=245), with relative sex proportions [21, 22]. **B.** Scatterplot and marginal densities of sequencing coverage and read length across all 493 individuals; HiFi and non-HiFi reads with read quality > 0.85 were combined. **C.** SVs linked to AD haplotypes. Each dot represents a SNP-SV pair: the x-axis shows the LD R^2^ calculated by scaling SV size to the [0, 2] interval; the y-axis shows the −log_10_(P) from a quantitative trait locus (QTL) model with SV size as the quantitative trait and the SNP as the predictor. **D.** Histogram of SV size, shown on a log10 scale. **E.** Distribution of major SV types, annotated with RepeatMasker. When multiple annotations overlapped an SV, the annotation with the largest overlap was retained. **F.** Distribution of major chromatin states overlapping SVs, based on 15-state ChromHMM annotations [26]. **G.** Fraction of biallelic (allele count ≤ 2) and multiallelic SVs, classified from the unique alleles observed across the 493 samples after joint genotyping. **H.** Genic context of SVs. **I.** Comparison of methylation-QTL (mQTL) strength: SNP-based mQTL −log_10_(P) on the y-axis versus SV-based mQTL −log_10_(P) on the x-axis. Asterisks (*) indicate significant differences between the full set and the linked subset by Fisher’s exact test.

Structural variant calling followed by joint genotyping identified 910,901 unique SVs genome-wide. After quality control, 20,483 variable SVs mapped within 250 kbp of the 280 AD-haplotypes (see Methods). The median size of this set of SVs in the reference genome GRCh38 was 50 bp (mean=89.7 bp, *Figure 2D*). These SVs were predominantly intronic (55%) or intergenic (40%; RefSeq annotations, *Figure 2H*) [23, 24], were frequently annotated as Alu transposable elements (54%) and tandem repeats (25%; RepeatMasker annotations, *Figure 2E*) [23, 25], and often overlapped transcription-associated (37%) or enhancer (10%) chromatin states (chromHMM annotations, *Figure 2F*) [26]. They were also highly polymorphic: 67% were multiallelic, with a median of five distinct allele sequences per locus across individuals (*Figure 2G*).

Together, these findings revealed extensive structural heterogeneity within AD-associated regions. In particular, the predominance of multiallelic SVs indicates that individuals assigned to the same biallelic SNP genotype may nevertheless carry several distinct structural alleles. We next tested which of these SVs were embedded within the independently associated AD-haplotypes.

### Most AD-haplotypes contain structural variation not revealed by the lead SNP

To determine how frequently AD-associated SNP markers tag underlying structural variation, and how completely they represent its allelic complexity, we quantified linkage between each AD-associated haplotype and nearby SVs. We combined (1) quantitative trait locus (QTL) analysis using SV length as a quantitative trait, with (2) conventional LD estimation based on SV allele lengths rescaled to the [0–2] interval to mimic SNP dosage (see Methods). QTL analysis naturally accommodates multiallelic SV variation, whereas conventional LD provides a more intuitive and widely used measure of genetic linkage. The two approaches showed near-complete concordance (Spearman’s ρ=0.99, *Figure 2C*), supporting the robustness of the inferred SNP-SV relationships.

Applying both the QTL association threshold (p<5×10^-8^) and the LD threshold (R^2^>0.15) identified 2,125 SNP-SV pairs, comprising 2,000 unique SVs across 207 of the 280 AD-associated haplotypes (74%, *Figure 2C* and *Table S4*). Some SVs were linked to multiple AD-associated SNPs, reflecting shared structural alleles across neighboring haplotypes, whereas for 73 haplotypes no SNP-SV pair showed both strong QTL-association and sufficient LD. Overall, 46 haplotypes contained at least one SV in high LD with the representative SNP (R^2^>0.8); 72 contained an SV in moderate LD (0.5<R^2^<0.8); and 89 contained SVs in lower LD (0.15<R^2^<0.5). Thus, while some structural alleles were almost completely represented by the lead SNP, many were only partially captured, indicating additional structural heterogeneity within the SNP-defined haplotypic background.

Compared with all 20,483 SVs located near AD-associated haplotypes, the 2,000 linked SVs were enriched in intronic (OR=1.18, p=8.9×10^-4^), enhancer (OR=1.22, p=9.1×10^-3^), and transcription-associated regions (OR=1.30, p=4.5×10^-8^), and depleted from heterochromatin (OR=0.76, p=9.7×10^-9^) and repressed chromatin states (OR=0.59, p=6.9×10^-4^) (*Figure 2E-H*) [25, 26]. They were also substantially more likely to be multiallelic than background SVs (OR=2.61, p=1.0×10^-61^). The median size of this refined set of SVs in GRCh38 was 52 bp (IQR=34, mean=89.3 bp). These findings indicate that the structural variation linked to AD-haplotypes is not a random sample of local genomic variation, but is preferentially associated with active regulatory contexts and with allelic complexity that cannot be represented fully by SNPs alone.

We next asked whether different SV alleles also co-occurred with distinct local epigenetic configurations. Significant associations between SV allele length and local methylation patterns were observed for 352 of the 2,000 SVs (18%, FDR<5%, *Table S5*). For 55% of the AD-haplotypes, the strongest SV-based methylation association exceeded the strongest nearby SNP-methylation association (paired Wilcoxon, p<2×10^-16^, *Figure 2I*). Moreover, AD-haplotypes carrying linked SVs were twice as likely to overlap with known expression- or splicing-QTL based on GTEx annotations (OR=2.01, p=0.01) [27].

Together, these results show that most AD-associated haplotypes contain structural and regulatory information that is incompletely represented by their lead SNP. In many cases, the linked SV showed a stronger association with local methylation patterns than the strongest nearby SNP, supporting the use of structurally resolved haplotypes as the relevant unit for functional interpretation.

### Regulatory SVs reveal several ways in which lead SNPs incompletely represent AD haplotypes

To prioritize SVs with potential regulatory relevance in AD-haplotypes, we integrated allele-specific differential methylation patterns, genomic localization, and chromatin-state annotation (see Methods). SVs significantly associated with local methylation, located within genes, and overlapping regulatory chromatin states (based on ChromHMM annotations) [26] were ranked highest and classified into three tiers of regulatory support (*Figure 3A*). After selecting one representative SV per AD-haplotype based on LD and functional prioritization (see Methods), 52 AD-haplotypes were characterized by a tier 1 SV (*Table 1*), 51 by a tier 2 SV, and 104 by a tier 3 SV (*Table S6* and *Table S7*). Among the 52 AD-haplotypes represented by a tier 1 SV, 14 showed strong LD between the representative SV and the corresponding lead GWAS SNP signal (LD R^2^ > 0.8), supporting its potential contribution to the underlying genetic association.

**Figure 3:**
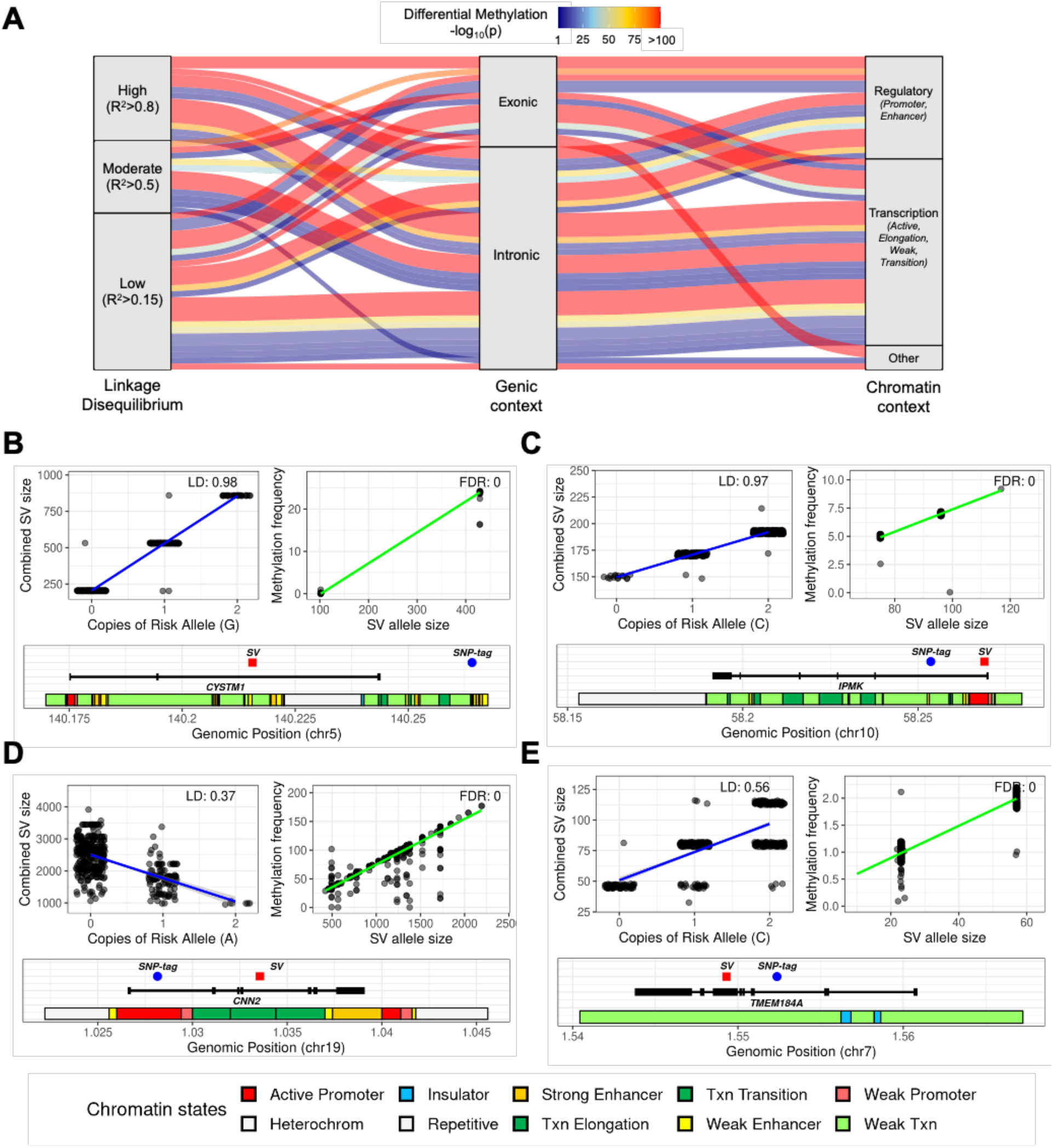
Prioritization of candidate regulatory SVs at AD loci. **A.** Alluvial plot of tier 1 candidate SVs, stratified by LD level with respect to the AD-haplotype, genic context, and chromatin context. **B.** Region surrounding *CYSTM1* gene, in which we identified an intronic AluYb8 insertion (red square, bottom plot) is in full LD with the intergenic AD-associated SNP (blue circle, bottom plot). The SNP risk allele (G) co-segregates with the expanded Alu allele (top left plot), which is associated with increased local methylation (top right plot). **C.** Region surrounding *IPMK* gene, in which an intronic SV is in high LD with the top AD-associated SNP. The SNP risk allele (C) co-segregates with the expanded SV allele (top left plot), which is associated with increased local methylation (top right plot). **D.** Region within *CNN2* gene, in which an intronic large tandem repeat is in low LD with the top SNP. The tandem repeat is highly multiallelic, and expanded alleles are associated with increased AD-risk and increased local methylation. **E.** Region within *TMEM184A*, in which an exonic SV in moderate LD with the top AD-associated SNP overlaps a transcriptionally active region. The SNP risk allele (C) co-segregates with the expanded version of the SV, which increases local methylation signal.

**Table 1:**
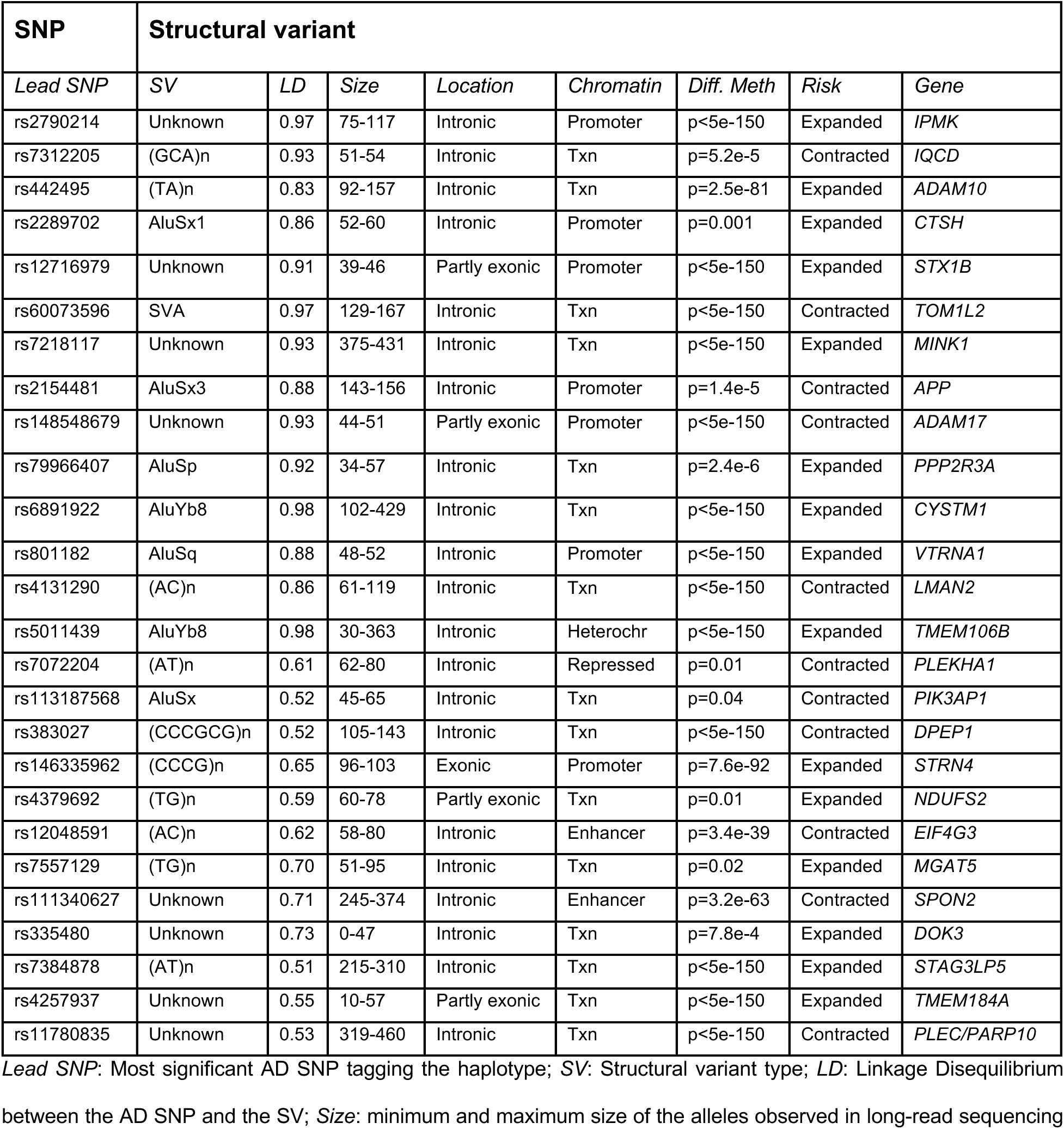
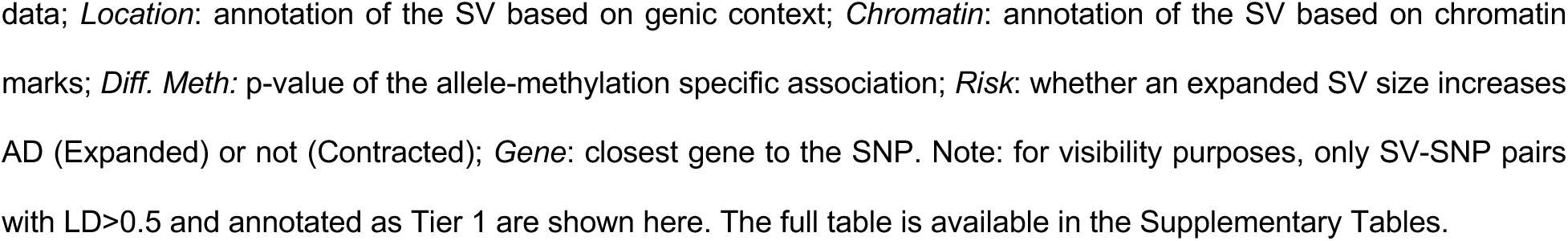
Tier 1 candidate regulatory SVs in moderate-to-high LD with AD haplotypes.

| SNP | Structural variant |  |  |  |  |  |  |  |
| --- | --- | --- | --- | --- | --- | --- | --- | --- |
| Lead SNP | SV | LD | Size | Location | Chromatin | Diff. Meth | Risk | Gene |
| rs2790214 | Unknown | 0.97 | 75-117 | Intronic | Promoter | p<5e-150 | Expanded | *IPMK* |
| rs7312205 | (GCA)<sub>n</sub> | 0.93 | 51-54 | Intronic | Txn | p=5.2e-5 | Contracted | *IQCD* |
| rs442495 | (TA)<sub>n</sub> | 0.83 | 92-157 | Intronic | Txn | p=2.5e-81 | Expanded | *ADAM10* |
| rs2289702 | AluSx1 | 0.86 | 52-60 | Intronic | Promoter | p=0.001 | Expanded | *CTSH* |
| rs12716979 | Unknown | 0.91 | 39-46 | Partly exonic | Promoter | p<5e-150 | Expanded | *STX1B* |
| rs60073596 | SVA | 0.97 | 129-167 | Intronic | Txn | p<5e-150 | Contracted | *TOM1L2* |
| rs7218117 | Unknown | 0.93 | 375-431 | Intronic | Txn | p<5e-150 | Expanded | *MINK1* |
| rs2154481 | AluSx3 | 0.88 | 143-156 | Intronic | Promoter | p=1.4e-5 | Contracted | *APP* |
| rs148548679 | Unknown | 0.93 | 44-51 | Partly exonic | Promoter | p<5e-150 | Contracted | *ADAM17* |
| rs79966407 | AluSp | 0.92 | 34-57 | Intronic | Txn | p=2.4e-6 | Expanded | *PPP2R3A* |
| rs6891922 | AluYb8 | 0.98 | 102-429 | Intronic | Txn | p<5e-150 | Expanded | *CYSTM1* |
| rs801182 | AluSq | 0.88 | 48-52 | Intronic | Promoter | p<5e-150 | Expanded | *VTRNA1* |
| rs4131290 | (AC)<sub>n</sub> | 0.86 | 61-119 | Intronic | Txn | p<5e-150 | Contracted | *LMAN2* |
| rs5011439 | AluYb8 | 0.98 | 30-363 | Intronic | Heterochr | p<5e-150 | Expanded | *TMEM106B* |
| rs7072204 | (AT)<sub>n</sub> | 0.61 | 62-80 | Intronic | Repressed | p=0.01 | Contracted | *PLEKHA1* |
| rs113187568 | AluSx | 0.52 | 45-65 | Intronic | Txn | p=0.04 | Contracted | *PIK3AP1* |
| rs383027 | (CCCGCG)<sub>n</sub> | 0.52 | 105-143 | Intronic | Txn | p<5e-150 | Contracted | *DPEP1* |
| rs146335962 | (CCCG)<sub>n</sub> | 0.65 | 96-103 | Exonic | Promoter | p=7.6e-92 | Expanded | *STRN4* |
| rs4379692 | (TG)<sub>n</sub> | 0.59 | 60-78 | Partly exonic | Txn | p=0.01 | Expanded | *NDUFS2* |
| rs12048591 | (AC)<sub>n</sub> | 0.62 | 58-80 | Intronic | Enhancer | p=3.4e-39 | Contracted | *EIF4G3* |
| rs7557129 | (TG)<sub>n</sub> | 0.70 | 51-95 | Intronic | Txn | p=0.02 | Expanded | *MGAT5* |
| rs111340627 | Unknown | 0.71 | 245-374 | Intronic | Enhancer | p=3.2e-63 | Contracted | *SPON2* |
| rs335480 | Unknown | 0.73 | 0-47 | Intronic | Txn | p=7.8e-4 | Expanded | *DOK3* |
| rs7384878 | (AT)<sub>n</sub> | 0.51 | 215-310 | Intronic | Txn | p<5e-150 | Expanded | *STAG3LP5* |
| rs4257937 | Unknown | 0.55 | 10-57 | Partly exonic | Txn | p<5e-150 | Expanded | *TMEM184A* |
| rs11780835 | Unknown | 0.53 | 319-460 | Intronic | Txn | p<5e-150 | Contracted | *PLEC/PARP10* |

**Lead SNP:** Most significant AD SNP tagging the haplotype; **SV:** Structural variant type; **LD:** Linkage Disequilibrium between the AD SNP and the SV; **Size:** minimum and maximum size of the alleles observed in long-read sequencing
data; *Location*: annotation of the SV based on genic context; *Chromatin*: annotation of the SV based on chromatin marks; *Diff. Meth*: p-value of the allele-methylation specific association; *Risk*: whether an expanded SV size increases AD (Expanded) or not (Contracted); *Gene*: closest gene to the SNP. Note: for visibility purposes, only SV-SNP pairs with LD>0.5 and annotated as Tier 1 are shown here. The full table is available in the Supplementary Tables.

### Lead SNPs as near-complete proxies for linked structural alleles

As expected, the previously described *TMEM106B* AluYb8 retrotransposon was among the strongest candidates. At the *CYSTM1* locus, the AD-associated haplotype tagged by rs6891922 (chr5:140,264,131, T>G, p_AD_=5.4×10^-7^) was in complete linkage (LD R^2^=0.98) with an intronic AluYb8 insertion located within a transcriptionally active region (*Figure 3B* and Supplementary results). The AD risk-increasing allele of the SNP (G) co-segregated with the expanded Alu allele, which was strongly associated with the presence of a distinct methylation pattern (FDR<2×10^-16^).

At the *IPMK* locus, the AD-associated haplotype tagged by rs2790214 (chr10:58,253,643, C>T; p_AD_ = 1.1×10^-9^) co-segregated with an intronic SV of 75-117 bp in an active promoter region (*Figure 3C* and Supplementary results). Larger SV alleles co-occurred with the AD risk haplotype (tagged by the SNP C allele) and showed marked allele-specific methylation patterns (FDR<2×10^-16^), consistent with differential regulation of the locus. Similar patterns were observed at additional loci including *LMAN2, CTSH, APP, TOM1L2,* and *MINK1*, where SVs in strong LD with AD-haplotypes localized within promoters or transcription-associated chromatin regions and showed evidence of allele-specific methylation patterns (see Supplementary results).

### Multiallelic SVs distinguish substructure within SNP-defined haplotypes

We next examined the 38 tier 1 SVs in moderate (LD 0.5<R^2^<0.8) or lower (LD 0.15<R^2^<0.5) linkage with AD-haplotypes. Although these variants do not fully co-segregate with the lead GWAS SNP, we identified strong methylation patterns for 12 SVs in moderate LD (*Table 1*) and 26 in low LD, indicating structural heterogeneity among chromosomes carrying the SNP-defined association signal. These SVs may therefore distinguish sub-haplotypic configurations that are not captured by the lead SNP alone (see Supplementary results). At *TMEM184A*, a partly exonic SV of 10-57 bp in moderate LD with rs4257937 (chr7:1,552,361, G>C) was associated with a pronounced methylation pattern, that distinguished between alleles (FDR<2×10^-16^, *Figure 3E*). At the *NDUFS2* and *STRN4* genes, the haplotypes tagged by variants rs4379692 (chr1:161,216,523, C>T) and rs146335962 (chr19:46,651,227, C>T) are both in moderate LD with exonic repeats: a (TG)n repeat ranging from 60-78 bp in size in *NDUFS2* (LD R^2^ = 0.59 with rs4379692); and a larger (CCCG)n repeat of size 96-103 bp in *STRN4* (LD R^2^ = 0.65 with rs146335962), localized within an active promoter and with expanded alleles significantly increasing methylation frequency (p<7.6×10^-92^). Similar regulatory repeat structures were observed at *EIF4G3, SPON2, MGAT5, CDK10,* and *PLEC*.

At *INPP5D*, a large AluSz insertion (293-443 bp) localized within transcriptionally active chromatin state and showed strong allele-specific methylation patterns (FDR<2×10^-16^) despite modest linkage (LD R^2^ = 0.28) with the AD-associated SNP rs13007032. Likewise, at *CNN2*, a highly multiallelic GC-rich tandem repeat of 500-2,250 bp overlapping an enhancer region was in weak linkage (LD R^2^ = 0.37) with the AD-haplotype and associated with strong methylation differences (FDR<5×10^-150^, *Figure 3D*). Similar configurations were observed at *ABCA1, TREML2, SLC24A4,* and *ZNF652* (see Supplementary results).

### Individual SVs intersect multiple independent association signals

Several loci also contained individual SVs linked to multiple independent AD-haplotypes. For example, an intronic SV of *ADAM10* (59-73 bp), overlapping an enhancer element showed allele-specific methylation patterns and low LD with the haplotypes tagged by rs2657125 (LD R^2^ = 0.22) and rs442495 (LD R^2^ = 0.18), which are independent of each other (LD R^2^ = 0.05). Similar examples included *CNN2, PLEC, PTK2B,* and *MS4A4A*, indicating that local structural variation can intersect with more than one SNP-defined association signal.

Together, these 52 candidates revealed that the relationship between representative GWAS SNPs and underlying structural variation is not one-to-one. At some loci, the SNP acted as a near-complete proxy for a linked SV; at others, the same SNP-defined signal encompassed multiple structural and epigenetic configurations; and in several regions, individual SVs occurred across more than one independently associated haplotypic background.

### SV-aware fine-mapping reveals multiple functional mechanisms at the *SHARPIN/PLEC* locus

We selected the *SHARPIN/PLEC* locus as a representative case study because it harbored three independent AD-associated haplotypes, including one linked to multiple tier 1-prioritized structural variants. We therefore asked whether resolving haplotype structure could refine the biological interpretation of this established GWAS locus. Previous GWAS identified two AD-associated missense variants in *SHARPIN*, rs34674752 and rs34173062 [3, 4, 28]. In addition to these previously reported signals, our conditional analysis resolved an independent AD-haplotype tagged by rs11780835*-G* (chr8:143987994; C:G), located ∼125 kb upstream of *SHARPIN* and <15 kb upstream of *PARP10* and *PLEC.* After conditional analysis, this haplotype remained associated with AD (GWAS p=2.95×10^-10^; COJO p=2.2×10^-3^, *Figure 4A*). Notably, the rs11780835*-G* risk allele is both an eQTL and splicing-QTL for *PLEC* across multiple brain regions (cerebellum, cortex, frontal cortex, hippocampus, hypothalamus, and cerebellar hemisphere, based on GTEx), consistently reducing gene expression [27]. Among genes within the locus, *PLEC* was the only one showing consistent brain eQTL and sQTL evidence, whereas *SHARPIN* showed no detectable brain QTLs for rs11780835. This is consistent with recent studies implicating reduced *PLEC* levels in AD-related glial activation, tau accumulation, and cognitive decline [29, 30].

**Figure 4.**
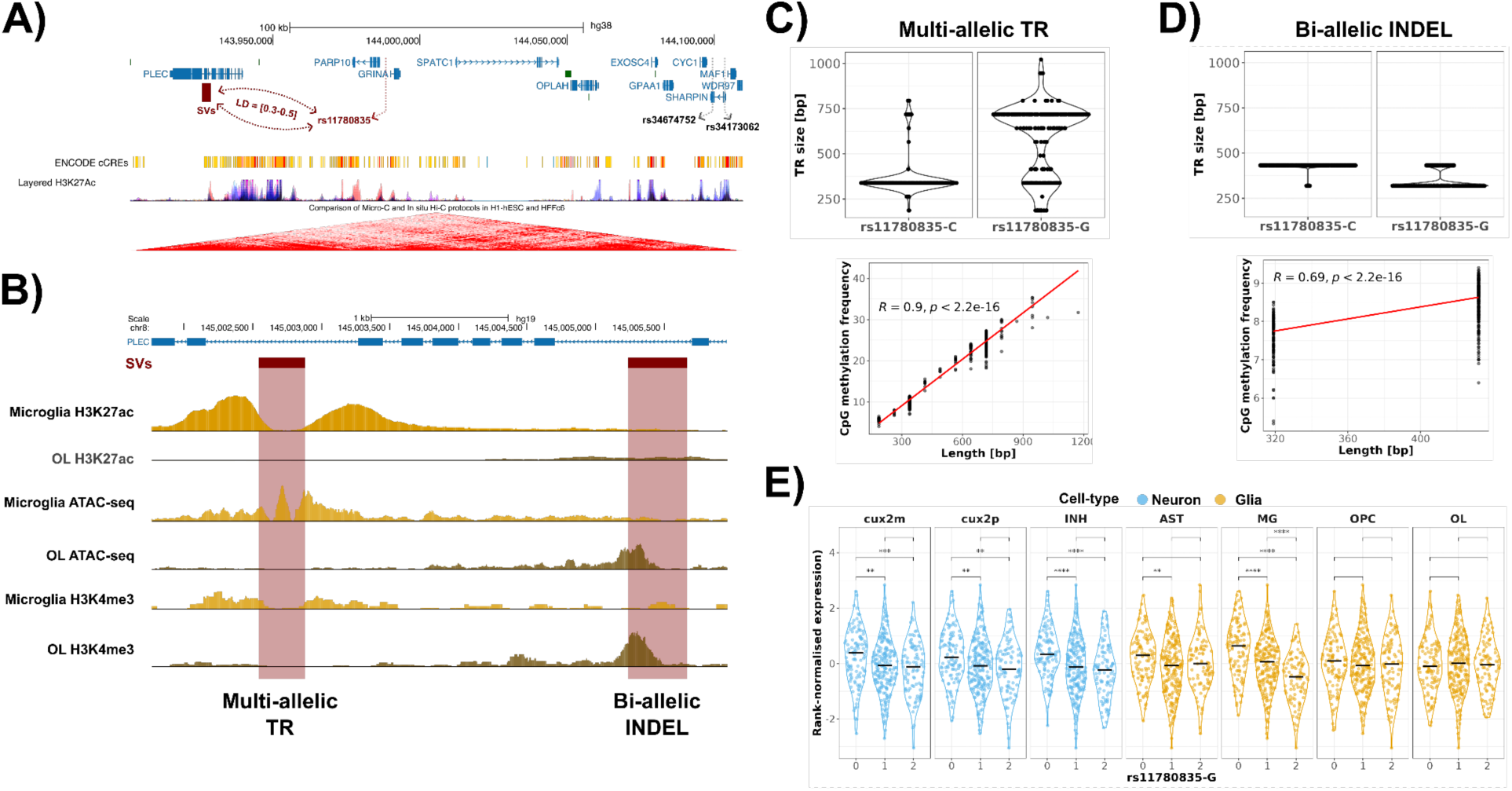
An AD-associated haplotype at the *SHARPIN/PLEC* locus is linked to candidate regulatory SVs in *PLEC*. **A.** UCSC genome browser view of the *SHARPIN/PLEC* locus, showing the two previously reported AD-associated missense variants in *SHARPIN* (rs34674752 and rs34173062), and the independent haplotype marked by rs11780835. The two tier 1 SVs with the strongest LD (R^2^=0.3-0.5) to rs11780835 are intronic in *PLEC*. **B.** Zoomed-in view of *PLEC* (top blue line, exons as blue rectangles), showing the intronic locations of the two SVs (maroon rectangles), with microglia and oligodendrocytes H3K27ac and ATAC-seq tracks indicating overlap with enhancer annotations (gold tracks). **C-D.** Top: allele-length distributions in rs11780835 allele carriers based on long-read phasing. Bottom: average methylation frequency (Y-axes) per allele sequence per genome (black dots) as a function of allele-length (X-axes). **E.** Single-cell eQTL analysis based on ROS/MAP data showing rank-normalised *PLEC* expression (Y-axis) as a function of rs11780835 allele dosage (X-axis) across cell-types (blue: neurons; yellow: glia).

Moving further to an SV-aware view, we observed that the rs11780835-haplotype comprised 15 linked SVs. These included (*i*) a biallelic intronic INDEL (LD R^2^ = 0.53) involving differential presence of 140 bp, where carriers of rs11780835*-G* frequently carried the deletion (*Figure 4D*); and (*ii*) a CpG-rich multi-allelic tandem repeat (LD R^2^ = 0.30) ranging from 0.18-1.1 kb, in which expanded alleles were associated with increased methylation (Spearman Correlation=0.9, p<2.2×10^-16^) and rs11780835*-G* preferentially harbored the expanded alleles (*Figure 4C*). Both SVs were prioritized as tier 1 candidates and overlapped enhancer annotations in microglia and oligodendrocytes, suggesting cell-type-specific regulation of *PLEC* (*Figure 4B*, see Methods). Although the regulatory impact of the INDEL remains unclear, the tandem repeat expansion may reduce *PLEC* expression through methylation-mediated enhancer inactivation, consistent with the observed eQTL effects. Supporting this model, single-cell eQTL analysis in the ROS/MAP cohort revealed significantly reduced *PLEC* expression in rs11780835*-G* carriers across cell types, with the strongest effect observed in microglia (−0.5 average change in rank-normalised expression; FDR-adjusted q <= 4.67×10^-6^, *Figure 4E*) [31].

Thus, resolving the locus beyond its previously reported lead variants revealed a more complex genetic architecture, comprising both coding *SHARPIN*-associated haplotypes and an independent regulatory *PLEC* haplotype involving structural variation, enhancer methylation and altered microglial expression. Functional experiments will be required to determine whether either *PLEC* SV is causal.

### Exploratory SV-aware models capture AD association structure beyond lead SNPs

We then assessed whether models representing multiple haplotypes and SVs captured local association structure more completely than single-lead-SNP models. We used directly genotyped SVs and local SNP haplotypes from 493 long-read-sequenced individuals to train imputation models, which were then applied to 2,381 AD cases and 2,293 healthy controls with available SNP genotyping data (*Figure 5A* and Methods). Of the 2,000 candidate SVs, 1,651 (83%) achieved good imputation quality (imputation R^2^>0.5), with a median imputation accuracy of 0.76 (*Figure 5B*, *Table S8*). Imputation performance increased with LD between the SV and nearby haplotype-tagging SNPs (Spearman’s correlation=0.28, p=2.5×10^-30^), as expected for SNP-based prediction. Using the imputed dataset, we tested both AD-haplotypes and SVs for association with AD status. Association testing replicated established AD loci including *BIN1, PLCG2,* and *PICALM* (FDR<0.10, *Figure 5C, Table S9*). In parallel, 112 imputed SVs showed significant association with AD risk (*Table S10*), including candidate regulatory SVs near *PICALM, NDUFAB1,* and *ACE* (FDR<0.10, *Figure 5C*).

**Figure 5:**
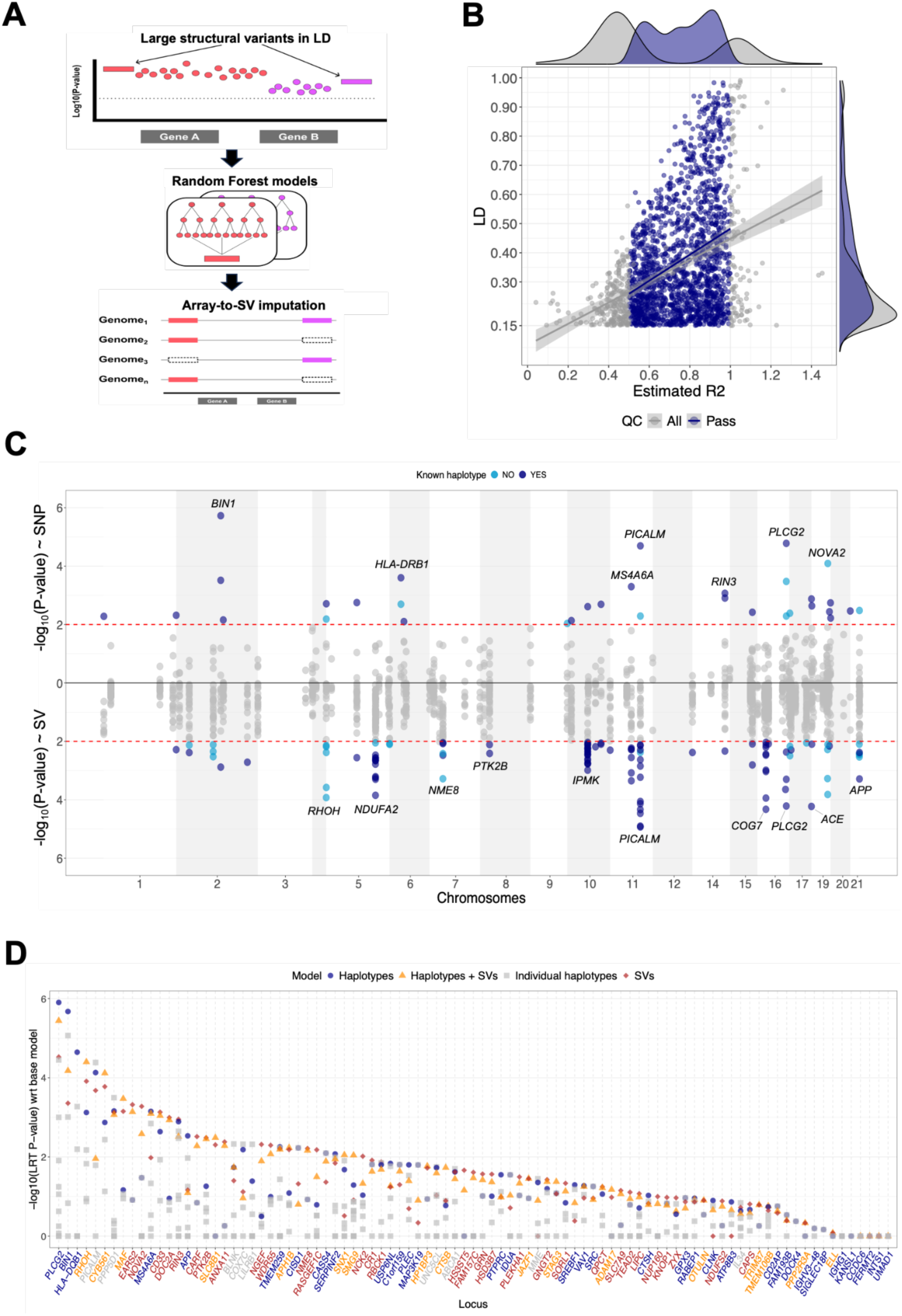
Imputation of SV sizes using SNP genotypes and association analyses. **A.** Schematic representation of the imputation workflow. Local SNP haplotypes were used to predict SV size. Random forest models were trained and evaluated by five-fold cross-validation within individuals with measured SV size from long-read sequencing data, and final models were applied to array-genotyped individuals. **B.** Correlation and marginal densities between imputation quality (R^2^) and LD between the SV and the AD-haplotypes, shown for all SVs and for the 1,651 SVs passing imputation QC (R^2^ > 0.5). **C.** Miami plot of association with AD for imputed SNPs and SVs, based on 2,381 AD cases and 2,293 controls. Significant associations after multiple-testing correction (FDR < 10%) are highlighted. Dark dots indicate SNPs and SVs in LD with known haplotypes; light dots indicate significant associations at additional haplotypes. Top genes are annotated for both SNPs and SVs. **D.** Comparison of association statistics between a baseline model including only PC1-5 and four extended models: (i) individual haplotypes (grey squares), (ii) combined haplotypes (blue circles), (iii) combined haplotypes plus SVs (orange triangles), and (iv) combined SVs (red diamonds). Models were fit using elastic-net regression combining L1 and L2 penalties, and compared with the baseline model as an exploratory ranking of local association structure.

We then assessed whether jointly modeling multiple haplotypes and SVs was preferred over single-SNP models. Model classes were ranked at each locus according to improvements in penalized model fit relative to simpler nested models, using the approximate likelihood-ratio procedure (see Methods). Across loci, the preferred model class varied, consistent with heterogeneous genetic architecture at AD-associated regions. Single-SNP models were favored in 10% of loci (e.g. in *CR1*); multiple-haplotypes models in 37% of loci (e.g., in *BIN1, PLCG2, HLA*, and *MS4A6A*); and SV-containing models in 53% of loci (*Figure 5D, Table S11*). Among loci showing nominal evidence over the baseline covariate-only model (LRT p<0.05), SV-containing models were favored in 33 of 56 loci (59%).

Because imputed SVs are partially inferred from local SNP haplotypes, and because penalized model comparisons are exploratory, these rankings should not be interpreted as evidence for statistically independent SV effects. Instead, they indicate that models incorporating structural variation or multiple haplotypes more often capture local association structure than lead-SNP-only models.

Together, these analyses suggest that AD-associated loci are frequently better represented by models incorporating multiple haplotypes and structural variation than by a single lead SNP alone. Although these comparisons are exploratory and do not establish independent SV effects, they indicate that structurally resolved haplotypes can capture aspects of local genetic architecture that are not fully represented by conventional lead-SNP models.

## Discussion

Genome-wide association studies have identified 98 risk loci for Alzheimer’s Disease [3, 4], but these associations are generally represented by individual lead SNPs while the underlying genetic architecture may be substantially more complex. Here, we show that the lead SNP is often an incomplete representation of the underlying haplotype structure at AD-associated loci. Across 98 established AD risk loci, we identified 280 conditionally independent association signals, indicating that many loci contain multiple independently associated haplotypes rather than a single SNP effect. Long-read whole-genome sequencing of 493 individuals further revealed 2,000 structural variants (SVs) linked to 207 of these 280 haplotypes. These SVs were enriched in intronic, enhancer and transcription-associated regions, were frequently multiallelic, and often co-occurred with distinct methylation patterns. Together, these findings show that lead GWAS SNPs frequently tag structurally and regulatorily heterogeneous haplotypes that cannot be fully understood through the SNP alone.

Integration of methylation, genomic context and chromatin annotation prioritized 52 AD-haplotypes with candidate regulatory SVs, illustrating how lead SNPs can incompletely represent the underlying haplotype structure. At loci such as *CYSTM1* and *IPMK*, the lead SNP acted as a near-complete proxy for an SV associated with a distinct methylation configuration. At other loci, including *CNN2*, *INPP5D* and *TMEM184A*, moderate or low linkage revealed structural heterogeneity within the SNP-defined association signal. Moreover, some SVs occurred across more than one independently associated AD-haplotype. Thus, the relationship between a GWAS lead SNP, the underlying AD-associated haplotype, and its structural alleles is often not one-to-one.

The *TMEM106B* locus provides an important proof of concept. Here, an AD-associated haplotype co-segregates with an intronic AluYb8 insertion associated with local methylation landscape and potential *TMEM106B* regulation [18, 32]. The present study extends this principle from a single locus to a systematic survey of AD-associated haplotypes. Many linked SVs were transposable-element insertions or tandem repeats, variant classes that are poorly represented by SNP arrays and difficult to resolve using short-read sequencing. Their recurrence across AD loci suggests that structurally complex alleles are an underappreciated component of the genetic architecture represented by GWAS associations.

Several prioritized loci also showed convergence with independent functional evidence. Candidate regulatory SVs at *PLEC, IPMK, LMAN2*, and *CNN2*, co-occurred with allele-specific methylation patterns and overlapped active enhancer or transcription-associated chromatin states. At *CTSH*, the haplotype tagged by rs2289702 has been experimentally linked to *CTSH* expression and microglial Aβ phagocytosis [33]. At *APP*, the rs2154481 haplotype acts as an eQTL with regulatory activity at an intronic enhancer while targeted methylation editing modifies *APP* expression and AD-relevant phenotypes in vivo [28, 34]. *IPMK* has similarly been associated with neurofibrillary tangle pathology and neuronal transcriptional regulation [35]. This convergence supports the biological relevance of the SV-aware prioritization, although it does not establish that the linked SVs themselves are causal.

The *SHARPIN/PLEC* locus provides the clearest example of how structural variation can refine interpretation of AD GWAS signals. Previous studies emphasized AD-associated missense variants in *SHARPIN*, whereas our conditional analysis identified an additional independent haplotype tagged by rs11780835-G and associated with reduced *PLEC* expression across multiple brain regions. Long-read fine-mapping revealed linked intronic SVs in *PLEC*, including a CpG-rich multiallelic tandem repeat whose length covaried almost perfectly with local methylation at an overlapping enhancer-associated chromatin region. The same haplotype was associated with *PLEC* expression in brain cells, with the strongest effect observed in microglia. This convergence of haplotype structure, repeat length, methylation and cell-type expression identifies an additional candidate regulatory mechanism involving *PLEC* that complements the previously recognized coding variants in *SHARPIN*. This is consistent with broader evidence that tandem repeat length variation can affect local methylation and gene expression [36, 37], and that plectin function is relevant to tau-microtubule dynamics and neuronal phenotypes [30].

Our results also have practical implications for GWAS fine-mapping. Even after resolving multiple independent association signals, interpreting each signal through a single lead SNP may overlook the underlying haplotype architecture and structural variation that contribute to disease risk. Using the long-read cohort as a reference panel, 83% of haplotype-linked SVs could be imputed into a larger SNP-genotyped cohort. In exploratory model comparisons based on penalized likelihood-ratio ranking, lead-SNP-only models were preferred at only 10% of loci, whereas models incorporating multiple haplotypes, SVs or both were preferred at the remaining 90%. Because the SVs were imputed from local SNP haplotypes, these analyses do not establish independent SV effects but they show that structurally resolved haplotype information can refine the representation of AD association signals at population scale.

Several limitations should be considered. The long-read discovery panel consisted of individuals of European ancestry, while SV frequencies, haplotype structures and imputation performances may differ across populations [38]. Methylation was measured in blood-derived DNA, whereas AD-relevant regulatory effects are likely to depend on brain region and cell type. Finally, methylation, chromatin annotation and expression-QTL evidence provide convergent prioritization but do not establish causality. Larger and more diverse long-read reference panels together with direct functional perturbation of prioritized SVs, will therefore be required.

Taken together, our findings show that AD GWAS associations are frequently embedded within structurally and regulatorily heterogeneous haplotypes that cannot be fully interpreted through the lead SNP alone. Rather than the lead SNP, the underlying haplotype should be regarded as the fundamental unit for resolving the biological mechanisms driving AD genetic risk.

## Methods

This section is organized around the same sequence of analyses used to address our central research question: we first identified conditionally independent AD-associated haplotypes from GWAS summary statistics, then identified structural variants linked to these haplotypes using long-read sequencing and evaluated their allele-specific methylation. Linked SVs were then prioritized into tiers of regulatory support based on methylation, genomic context and chromatin annotation, and the SHARPIN/PLEC locus was examined in depth as a representative case study of this prioritization. We then compared SNP-based, haplotype-based and SV-aware association models to assess whether they captured AD association structure more completely than the lead SNP alone. Finally, we evaluated whether these structurally resolved haplotypes could be imputed into larger SNP-genotyped cohorts to improve representation of AD association signals at population scale.

### Genotyping data

We included 4,674 genetically unrelated individuals of European ancestry: (i) 443 cognitively healthy centenarians from the 100-plus Study [22]; (ii) 88 healthy individuals from the 100-plus Study, corresponding to the partners of the centenarians’ offspring [22]; (iii) 1,951 patients clinically diagnosed with probable or possible AD from the Amsterdam Dementia Cohort (ADC) [21]; (iv) 430 pathologically confirmed AD patients from the Netherlands Brain Bank (NBB) [39]; (v) 1,612 Dutch healthy individuals from the Longitudinal Aging Study of Amsterdam (LASA) [40, 41]; (vi) 54 controls from the NBB [39]; (vii) 96 individuals from the Dutch twin registry [42]. Detailed description of the individuals included, their respective cohorts, and the quality control of the genetics data has been reported previously [43]. All individuals were genotyped with the Illumina Global Screening Array. Standard quality control methods were applied: individual call rate >99%, variant call rate >99%, Hardy-Weinberg equilibrium p-value<1×10^-6^. Genotypes were imputed with TOPMed, and variants with imputation quality R^2^>0.2 were retained for downstream analyses.

### Selection and association of AD variants from GWAS

We selected N=118 lead SNPs identified to have a genome-wide significant association with AD from previous AD-GWAS (with coordinates aligned to GRCh38) [4]. SNPs associated with *APOE,* rs7412 and rs429358 were excluded. For each lead SNP, we extracted GWAS summary statistics for all SNPs within 250 kbp (kilo base pairs). Pairwise linkage disequilibrium (LD) among SNPs was estimated in the full genotyped cohort of 4,674 individuals. We first performed clumping with plink2 (v2.00a6LM) [44] using an LD R^2^>0.15, a window of 250 kbp, and a suggestive association threshold of p<5×10^-5^. This step retained common haplotypes with suggestive evidence of association with AD, each represented by the most strongly associated SNP in the clump.

We then refined these signals with GCTA-COJO (v1.94.1) [45], using the stepwise selection model (--cojo-slct), a minor allele frequency threshold of >2.5%, and a joint-effect p-value of association p<0.05. Associations were corrected for multiple testing (False Discovery Rate, FDR), and SNPs with a conditional FDR<1% were considered significant. The resulting SNPs were used as representatives of conditionally independent candidate AD-haplotypes. Throughout this study, the term AD-haplotype therefore denotes the SNP-defined haplotypic background tagged by each conditionally independent representative SNP, rather than a haplotype reconstructed directly from phased long-read sequencing data.

### Long-read sequencing data and processing

Long-read whole-genome sequencing data were available for 493 individuals sequenced on the PacBio Sequel II platform. This discovery panel included 245 AD patients from the Amsterdam University Medical Center, and 248 cognitively healthy centenarians from the 100-plus Study cohort. Additional information regarding the samples included, the respective cohorts, and the long-read sequencing data generation and quality assessment is available elsewhere [18, 38, 43]. Genomic DNA was extracted from whole blood and assessed for integrity before sequencing [18, 38]. Raw reads were processed with an in-house pipeline (https://github.com/holstegelab/snakemake_pipeline), which generated high-quality HiFi reads and lower-quality non-HiFi reads with read quality >0.85. Reads were aligned to GRCh38 patch release 14 with pbmm2, a PacBio wrapper of minimap2 [46]. For samples sequenced across multiple SMRT cells, per-cell BAM files were merged into a single per-sample BAM file. HiFi and non-HiFi reads were combined for downstream structural-variant analysis to improve representation of larger and more complex alleles.

The Medical Ethics Committee (METC) of the Amsterdam UMC approved all studies contributing genotype or long-read sequencing data. All individuals and/or their legal representatives provided written informed consent for participating in clinical and genetic research.

### Structural variant calling and repeat annotation

Structural variants (SVs) were identified as described previously [18]. Candidate SVs were detected with sniffles2 (v2.0.7) [47] using default parameters. SV annotations, including simple repeats, satellites, and low complexity regions, were obtained from the UCSC Genome Browser [23], and intersected with candidate SV coordinates. To improve genotyping of structurally complex loci, local assembly and joint genotyping were performed with Otter (v1.0) [38]. Because HiFi-only analyses can be susceptible to allele dropout events, particularly for larger SVs, local assembly used both HiFi and non-HiFi reads with read quality >0.85 [38]. This procedure generated a joint-genotyped SV gVCF across all long-read samples. SVs overlapping centromeric regions or known segmental duplications, based on UCSC annotations, were excluded. For analyses of AD loci, we retained SVs located within 250 kb of AD-associated GWAS loci. Additional filters removed SVs genotyped in fewer than 10% of individuals, SVs with a local-to-global coverage ratio <0.3 or >3.3 [38], and SV alleles supported by fewer than three sequencing reads per allele.

### SNP-SV linkage analysis

We identified SVs linked to AD-associated haplotypes by combining a quantitative-trait-locus (QTL) framework with conventional linkage disequilibrium (LD) estimation. Because many SVs were multiallelic, SV allele length was modeled as a quantitative phenotype. For each individual, the phenotype was defined as the summed length of the two observed SV alleles. SNP dosage was tested for association with SV length by linear regression in plink2 (v2.00a6LM) [44], adjusting for the first five genetic principal components. For each SV, all SNPs within 250 kb were tested.

Conventional SNP-SV LD was estimated by linearly scaling SV lengths to the interval [0, 2], approximating diploid dosage values, and calculating LD in plink2. Concordance between the QTL and LD approaches was assessed by correlating QTL significance (-log_10_P) with LD estimates. SNP-SV pairs were retained when they met both a QTL threshold of p<5×10^-8^ and a minimum LD threshold of R^2^>0.15. Retained SNP-SV relationships were intersected with the set of conditionally independent AD-associated haplotypes identified by GCTA-COJO [45].

LD categories were defined as mutually exclusive bins: high LD, R^2^>0.8; moderate LD, 0.5<R^2^<0.8; and low LD, 0.15<R^2^<0.5.

### Methylation analysis

Allele-specific methylation profiles were derived from long-read BAM files with haplr (https://github.com/holstegelab/haplr), a probabilistic framework that assigns long-reads from BAM files to haplotypes using a Bayesian model based on haplotype-informative variants. For each read, haplr calculates posterior probabilities across competing haplotypes by integrating allele matches at informative sites while accounting for sequencing-error probabilities. Reads meeting predefined confidence criteria were assigned to the most likely haplotype. Methylation calls embedded in the BAM files were then aggregated by haplotype to obtain haplotype-specific CpG coverage, methylation counts, and methylation fractions. To test whether SV allele size was associated with allele-specific methylation, we modeled methylation as a function of SV allele size using linear regression. The model related haplotype-specific methylation levels to SV allele size. P-values were corrected for multiple testing using False Discovery Rate (FDR) and considered significant at FDR<5%.

We also performed cis-methylation quantitative trait locus (mQTL) analyses to compare SVs and SNPs as predictors of nearby CpG methylation. For each SV, CpG sites located within ±250 kb were considered if methylation measurements were available in at least 25% of individuals. CpG methylation was tested for association with SV length or SNP genotype dosage using robust linear regression models with the *rlm* function in the MASS package, adjusting for the first five genetic principal components. Associations were considered significant at FDR<5% across all SV-CpG and SNP-CpG tests. For loci in which SNPs and SVs were in LD, we directly compared the strength of methylation association signals between variant classes using paired Wilcoxon test on −log_10_(P) values.

### Prioritization of SVs

SVs were prioritized with a composite functional annotation score that integrated methylation evidence, genic localization and chromatin context. Four components were scored: differential methylation status, genic context, genic annotation and chromatin state. Lower scores indicated higher regulatory priority. Differential methylation was scored as 1 for FDR-adjusted P < 0.05, 2 for FDR-adjusted P ≥ 0.05 and 3 for missing methylation data. Genic context was scored as 1 for genic, 2 for intergenic and 3 for other or missing annotations. Genic annotation was scored as 1 for fully exonic, 2 for partly exonic, 3 for intronic and 4 for missing annotation. Chromatin state was scored as 1 for enhancer or promoter, 2 for transcription-associated, insulator or repressed states and 3 for other states. The methylation component was weighted twofold to emphasize direct epigenetic evidence. The final composite score was computed as the sum of the four components. Tier boundaries were defined from the empirical score distribution, which ranged from 5 to 15, with median 12 and mean 11.7. Tier 1 was defined as scores ≤ 8, tier 2 as scores 9-10, and tier 3 as scores > 10. To obtain a single representative SV for each AD-haplotype, SNP-SV pairs were first stratified by LD (high: R^2^>0.8; moderate: 0.5<R^2^<0.8; low: 0.15<R^2^<0.5). For each AD-haplotype, the representative SV was selected from the highest LD category available. Within that category, the SV with the lowest composite score was retained. Thus, LD with the GWAS SNP was prioritized over the composite score when selecting representative SVs. Tier 1 SVs were considered candidate regulatory SVs for follow-up, not causal variants.

### Fine-mapping analysis of the *SHARPIN/PLEC* locus

At the *SHARPIN/PLEC* locus, we evaluated SVs that were ≥ 25 bp and were at least in low LD (R^2^≥0.15) with rs11780835. Independence of the rs11780835 haplotype from the two previously reported *SHARPIN* missense variants (rs34674752 and rs34173062) was established during the genome-wide conditional analysis described above, in which all candidate signals within the locus were jointly modeled by GCTA-COJO. Among the linked SVs, two passed the tier 1 regulatory-candidate filters, and both were located within introns of *PLEC*. To link rs11780835-G carriership to SV alleles, we performed haplotype-resolved phasing of long reads with haplr. For each long-read genome, reads were assigned to one of two haplotypes using spanning alignments at nearby heterozygous common SNPs, and rs11780835 allele status was determined per haplotype. This yielded haplotype-resolved SV length for each rs11780835-allele. Using the same haplotype-resolved read sets, we then quantified allele-specific methylation. For each haplotype, CpG sites within the SV-spanning sequence were extracted and the average number of methylated CpGs was calculated. A CpG was considered methylated when the base-modification probability encoded in the BAM ML tag was at least 50%.

To compare SV locations with cell-type-specific enhancer annotations, SV coordinates were lifted over from GRCh38 to GRCh37 with the UCSC Genome Browser and overlapped with H3K27ac and ATAC-seq peaks from Nott et al. [48]. Cell-type-specific eQTL analysis tested the association between rs11780835 allele dosage and reduced *PLEC* expression in single-nucleus RNA-sequencing data from ROS/MAP cohort, following the procedure described in Salazar et al. [18], and based on the d-sum normalization procedure of Cuomo et al. [49].

### SV imputation from SNP genotypes

For each SV, we trained a random-forest regression model to predict SV size from nearby SNP genotypes. Predictor SNPs were selected from variants in LD with the top SNP of the corresponding AD-associated haplotype (R^2^ > 0.15). Model tuning used fivefold cross-validation within the long-read cohort, with up to 493 measured samples per SV depending on missingness. A final model was then trained on all long-read samples with measured SV genotypes for that SV and applied to the 4,674 SNP-array-genotyped individuals to impute SV size. Imputation quality was summarized with an R^2^ metric analogous to the BEAGLE dosage R^2^ [50], defined as the ratio of the expected variance of imputed SV sizes in the training set to the observed variance of imputed SV sizes in the 4,674-sample target set. This R^2^ is an imputation-quality metric and should be distinguished from cross-validated predictive accuracy within the long-read cohort. SVs with imputation R^2^ > 0.5 were retained for downstream analyses.

After imputation, we performed a SV-GWAS analysis by comparing the size of each SV between 2,381 AD cases and 2,293 healthy controls. In the same sample set, we also performed a SNP-GWAS analysis by comparing the dosages of AD-haplotypes. In both cases, we used logistic regression models (implemented in R, Wald test), correcting for the first five genetic principal components. Association p-values were corrected using FDR, and the same correction factor was applied to both SNP- and SV-GWAS associations.

### Comparison of SNP-based, haplotype-based and SV-aware association models

We evaluated whether models that include multiple AD-associated haplotypes and linked SVs better captured local AD association structure than models containing individual haplotypes (i.e., SNPs) alone. For each AD-associated locus, the COJO analysis defined n_haplo_ independent, significant haplotypes, each represented by its top SNP. For the same locus, SNP-SV linkage analysis identified n_SV_. Using 2,381 AD cases and 2,293 healthy controls, we first fit a baseline logistic model containing only the first five principal genetic components. We then evaluated four extensions: (1) an individual-haplotype model, fit separately for each haplotype and summarized by the best-performing single-haplotype fit per locus; (2) a multiple-haplotype model, containing all locus haplotypes; (3) an SV-only model containing linked SVs; and (4) a combined model containing both haplotypes and linked SVs.

Models were fit with logistic elastic-net regression, combining L1 and L2 penalties to reduce collinearity among predictors and encourage sparsity. The regularization mixing parameter α and penalty strength λ were selected by cross-validation. Because standard likelihood-ratio test assumptions do not hold for penalized elastic-net fits, we used an LRT-style comparison only as an approximate model-ranking procedure. Residual deviance from the H2O training output [51] was used as a proxy for the log-likelihood, and the number of non-zero coefficients in each model was used as the effective degrees of freedom (df). The resulting test statistic was evaluated against a χ^2^ distribution with degrees of freedom equal to the difference in non-zero coefficients between nested models. These approximate p-values were used to rank nested models and should not be interpreted as formal null-hypothesis tests. Where unpenalized logistic regression comparisons were applied, standard likelihood-ratio tests were used. As a sensitivity analysis, we evaluated model stability with k-fold cross-validation by comparing training AUC with cross-validated out-of-fold AUC. The maximum observed difference was 4.8 percentage points, providing limited evidence of overfitting under this assessment.

## Data availability

Individual-level human genetic and methylation data generated in this study will be made available in accordance with participant consent and institutional and data-access approvals through an appropriate controlled-access repository or, where deposition is not permitted, through a clearly described access procedure for qualified researchers.

Derived, non-identifying resources generated by the study, including haplotype-linked SV annotations, SV summary results and imputation-related resources are available in Supplementary Tables.

## Code availability

All analyses were performed with publicly available software, used as described in the Methods: sniffles2 (v2.0.7), TREAT/Otter (v1.0), plink2 (v2.00a6LM), GCTA-COJO (v1.94.1), pbmm2, and R packages MASS and h2o. Custom code developed for this study is available under an open-source licence at https://github.com/holstegelab/snakemake_pipeline (long-read sequencing data processing), https://github.com/holstegelab/haplr (haplotype-resolved read assignment and allele-specific methylation), and https://github.com/holstegelab/GWASum (identification of significant and independent AD haplotypes from GWAS summary statistics).

## Acknowledgements

We thank all participants of the 100-plus Study, the Amsterdam Dementia Cohort, the Longitudinal Aging Study Amsterdam, the Netherlands Brain Bank and the Netherlands Twin Register for their contribution to this research, as well as their families and caregivers. We are grateful to the ROS/MAP investigators for making single-nucleus expression data available. This work was supported by Scientific Excellence program 2014 (IPB), Stichting Alzheimer Nederland (WE09.2014-03), Stichting Dioraphte (VSM 14 04 14 02) and Stichting VUmc Fonds, Hans und Ilse Breuer Stiftung Alzheimer Research Prize 2020. H. Holstege and M.J.T. Reinders are recipients of ABOARD, a public-private partnership receiving funding from ZonMW Nationaal Dementiaprogramma (#73305095007) and Health∼Holland, Topsector Life Sciences & Health (PPP-allowance; #LSHM20106). More than 30 partners, including de Hersenstichting (Dutch Brain Foundation) participate in ABOARD (https://www.alzheimer-nederland.nl/onderzoek/projecten/aboard).

## Author contributions

N.T., A.S., G.B., and H.H. conceived and designed the study. N.T., A.S., and G.B. contributed equally and performed the primary analyses, with contributions from D.A., Y.Z., M.Hul, and S.v.d.L. N.T. and A.S. developed the analysis pipelines and performed the long-read sequencing analyses. L.K., S.W., and J.K. generated the long-read sequencing data and performed sample preparation and quality assessment. N.M.v.S. and M.Hui. contributed cohort data and phenotypic information from LASA. B.M.T., E.G.B.V., and S.v.d.L. contributed clinical and cohort data from the Amsterdam Dementia Cohort. H.H. contributed cohort data from the 100-plus Study. M.R., S.v.d.L., and H.H. supervised the study and acquired funding. N.T., A.S., and H.H. wrote the manuscript. All authors read, revised, and approved the final manuscript.

